# Impact of the Covid-19 Pandemic on Audiology Service Delivery: Observational Study of the Role of Social Media in Patient Communication

**DOI:** 10.1101/2023.06.23.23291820

**Authors:** Adeel Hussain, Zain Hussain, Mandar Gogate, Kia Dashtipour, Adele Goman, Aziz Sheikh, Amir Hussain

**Affiliations:** School of Computing, Edinburgh Napier University, Edinburgh, EH10 5DT, Scotland; The University of Edinburgh, Chancellor’s Building, 49 Little France Crescent, Edinburgh, EH16 4SB; Usher Institute,The University of Edinburgh, Old Medical School, Teviot Place, Edinburgh, EH8 9AGA, Scotland

## Abstract

The Covid-19 pandemic has highlighted an era in hearing health care that necessitates a comprehensive rethinking of audiology service delivery. There has been a significant increase in the number of individuals with hearing loss who seek information online. An estimated 430 million individuals worldwide suffer from hearing loss, including 11 million in the United Kingdom. The objective of this study was to identify NHS audiology service social media posts and understand how they were used to communicate service changes within audiology departments at the onset of the Covid-19 pandemic.Facebook and Twitter posts relating to audiology were extracted over a six week period (March 23 to April 30 2020) from the United Kingdom. We manually filtered the posts to remove those not directly linked to NHS audiology service communication. The extracted data was then geospatially mapped, and themes of interest were identified via a manual review. We also calculated interactions (likes, shares, comments) per post to determine the posts’ efficacy. A total of 981 Facebook and 291 Twitter posts were initially mined using our keywords, and following filtration, 174 posts related to NHS audiology change of service were included for analysis. The results were then analysed geographically, along with an assessment of the interactions within the included posts. NHS Trusts and Boards should consider incorporating and promoting social media to communicate service changes. Users would be notified of service modifications in real-time, and different modalities could be used (e.g. videos), resulting in a more efficient service.

## Introduction

To mitigate the exponential spread of Covid-19 cases, the government took quick and decisive steps that affected the healthcare services, including audiology departments in the UK. Consequently, there was a transition from in-person to teleaudiology patient care, which resulted in the cancellation of all in-person appointments for routine services. Moreover, new assessments and existing hearing aid users were no longer able to access their regular care pathways in the event of hearing difficulties [1, 2].

Prior to the service being shifted to remote or postal services, audiology was predominantly a face-to-face facility for patients. However, due to the lack of teleaudiology infrastructure, certain procedures, such as otoscopy, the necessity for sound-treated rooms for testing, and multiple face-to-face appointments for hearing aid fitting, counselling, and troubleshooting could not be performed remotely. Previous research [2, 3] has shown some reticence towards teleaudiology being used in clinical settings. Eikelboom and Swanepoel [3], surveyed 269 audiologists from around the world and found that only 15% had used teleaudiology, despite being confident in using the required technology [3]. Similarly, Saunders and Roughley surveyed 120 UK-based audiologists during the Covid-19 pandemic and found that while 98% of participants used teleaudiology at the time of the study, only 30% had used it beforehand. Participants responded positively to the remote pathways, but also highlighted the need for improvements in training and infrastructure [4]. In addition, the lack of infrastructure and the inability to perform certain tasks, teleaudiology services had to be utilised during national and regional lockdowns. In recent decades, patients are increasingly seeking more information, making informed rehabilitation decisions, and taking responsibility for managing their conditions. Previously, individuals with a disability or health issue would seek advice and assistance from the internet, their friends, family, or healthcare experts. With improved internet accessibility and social media, individuals have more autonomy in gathering information from a wide range of resources [5]. During the Covid-19 pandemic, face-to-face communication with healthcare providers decreased, which has exacerbated this trend. In April 2020, research studies showed that people in the United Kingdom spent an average of four hours a day online, up from three hours and 29 minutes in September 2019, further supporting the aforementioned [6]. A recent study by Gupta et al [7] found Facebook and Twitter to be the most commonly used type of social media. Approximately 430 million people worldwide and 11 million individuals in the United Kingdom are estimated to have some degree of hearing loss, approximately, 1 in 6 around 900,000 of them having severe or profound hearing loss [8]. A listener’s level of difficulty will vary based on the type of hearing loss they have, the environment they are in, whether their hearing loss has been treated, and how effectively their hearing aids are functioning. Since face-to-face communication was restricted due to the lockdown, a written source of information would have been beneficial if a listener had non-functioning hearing aids who were unable to participate in a telephone discussion. Our study is the first of its kind to investigate the role and impact of social media in engaging audiology consumers on service changes during the initial 6 weeks of the Covid-19 pandemic. Specifically, we aim to analyse how audiology departments across health trusts in the United Kingdom utilised Facebook and Twitter, to communicate with their patients.

## Materials and methods

### Ethics

Due to the availability of the data used in this study in the public domain, no NHS ethics review was required. Prior to previous research and to ensure compliance with pertinent provisions of the General Data Protection Regulation (GDPR) [9, 10], a comprehensive assessment was conducted to confirm that our study presented no privacy risk to individuals. We aimed to follow best practices for user privacy by excluding private information from our dataset. In addition, to comply with privacy laws and social media policies in accordance with the GDPR, to collect data, we did not disclose or published direct posts by individuals, quotations from individuals, or the names or locations of users who are not public organisations or entities on the Facebook CrowdTangle platform and Twitter API [11].

### Data sources

We opted to use data from Facebook and Twitter, since they are the most used social media platforms [7]. Within the period from March 23 to April 30, 2020, we specifically targeted English-language Facebook posts from pages in the UK. This particular time frame was selected due to its alignment with the initial announcement of the first UK lockdown [9]. In order to identify which audiology departments were utilising social media to communicate service changes, we narrowed our search to only include the initial six-week period of the lockdown. Furthermore, to collect Facebook posts and tweets, we employed the Crowdtangle service and Twitter API [12].

The search terms used to extract data were hearing loss, hearing, difficulty, presbycusis, tinnitus, deafness, speech impairment, hearing aids, audiology, ear wax, ear syringing, microsuction, telecare, teleaudiology remote consultations. The comprehensive range of our search terms ensured that all relevant results were included during the searches. Our initial search yielded 981 Facebook posts and 291 Twitter posts, which were then filtered to exclude posts unrelated to the NHS audiology departments. Our team manually assessed each post for messages of service change from NHS Trusts or partners, such as libraries where repair/battery clinics had previously been held. The final dataset did not include posts about private hearing clinics, other hearing-related news, or information from news outlets. This study also omitted information from the private sector and hearing aid manufacturers.

### Data analysis

Crowd tangle and Twitter API was used to calculate the frequencies of the various interactions from the filtered dataset. For parameters of interest, frequencies were calculated, and circular charts, tables, and heat maps illustrated the results. The presented heat map depicts the diverse geographical origins of the posts, with the colour and size of the circles serving as indicators of the quantity of interactions.

## Results and Discussion

In this study, we investigated how NHS audiology departments utilised social media to provide service updates to patients. Fig 1 illustrates that most posts were generated between the third and the fifth weeks.

**Fig 1.**
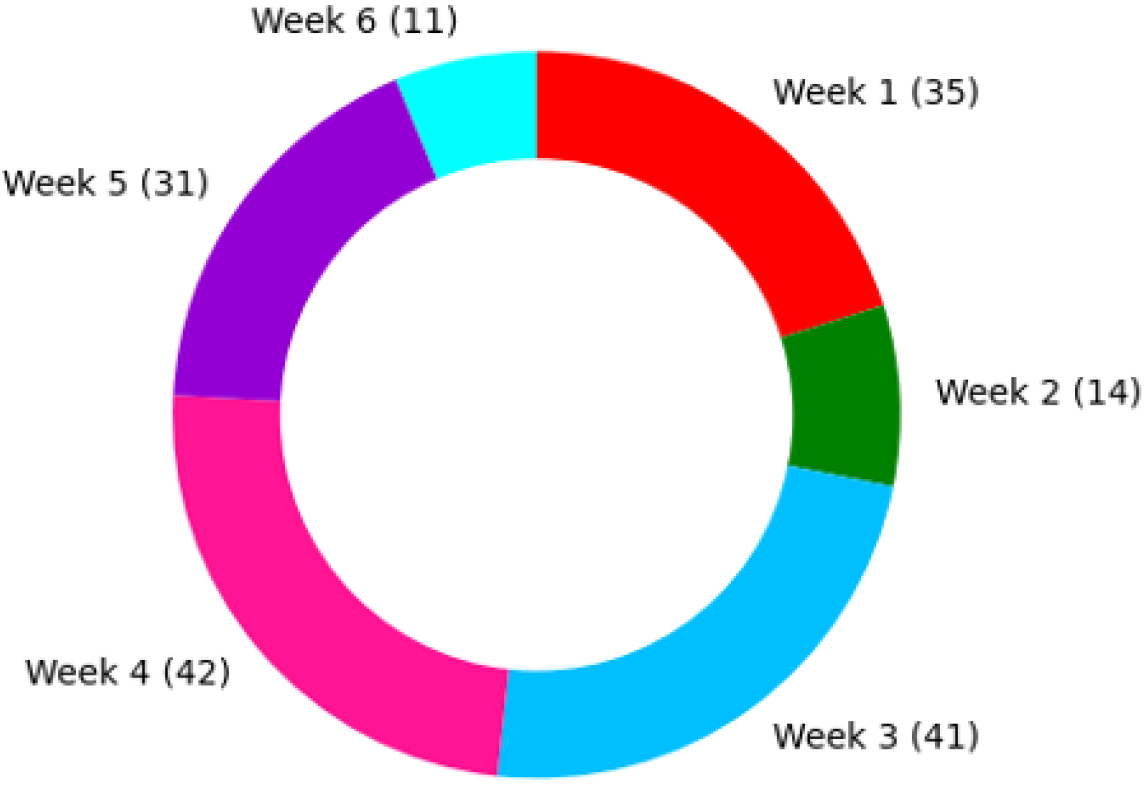
Number of weekly posts

Table 1 shows that out of 14 NHS Boards in Scotland, seven reported changes in service information on Facebook. Similarly, out of 217 NHS trusts in England, 41 reported changes in service information on Facebook and Twitter. In Wales out of 7 trusts, three reported a change in service information, and in Northern Ireland, out of five trusts only one reported a change in service information.

**Table 1.** Number of Trusts and Boards in different parts of the United Kingdom.

| Area | Number of NHS trusts boards | Number of trusts boards with posts of service change |
| --- | --- | --- |
| England | 217 | 41 |
| Scotland | 14 | 7 |
| Wales | 7 | 3 |
| Northern Ireland | 5 | 1 |

There was a total of 3,646 interactions across 174 posts, Table 2 illustrates the responses and interactions from the different countries across the UK. To be noted, this sample represents only a fraction of 11 million hearing aid users in the UK [8]. Interactions included likes, shares, comments, or reposts. However, it’s possible that some people who viewed the posts did not interact with them.

**Table 2.** Number of shares, likes and comments on posts in England, Scotland, Wales and N. Ireland

| Country | Likes | Shares | Comments | CTotal |
| --- | --- | --- | --- | --- |
| England | 915 | 856 | 236 | 2007 |
| Scotland | 415 | 737 | 323 | 1475 |
| Wales | 84 | 161 | 29 | 274 |
| Northern Ireland | 66 | 14 | 4 | 84 |

Fig 2 shows a visual representation of the interactions. Similarly, the bubble map indicates that the areas with a higher number of responses corresponded to areas with a higher number of interactions. In addition, the number of responses from the areas that posted service change updates in the U.K. Both the gradient of the filled circle and the size of the circle indicate the number of posts and the number of interactions per post using the Virdis scale (blue-¿yellow and small-¿large indicating an increasing number of interactions It has been shown that individuals who have hearing loss can benefit from utilising electronic media as it improves their communication abilities and reduces auditory barriers [13]. Therefore, internet usage appeals to those who prefer text-based communication [14]. It is recommended to leverage the impact of social media for obtaining service change updates, given that a study by Manchaiah et al [5] found that Facebook and YouTube were the most frequently used social media platforms, with over 40% of participants reporting using them in their study [5]. The current study provides evidence that sharing service change messages on reliable online platforms can effectively inform patients. However, the credibility of information shared on social media can be undermined by the circulation of false or misleading content, as indicated by previous research [7]. The failure to timely post accurate information on social media may result from the ambiguity surrounding lockdown protocols and a shortage of personnel due to the government furlough programme, which was initiated on 27 March 2020.

**Fig 2.**
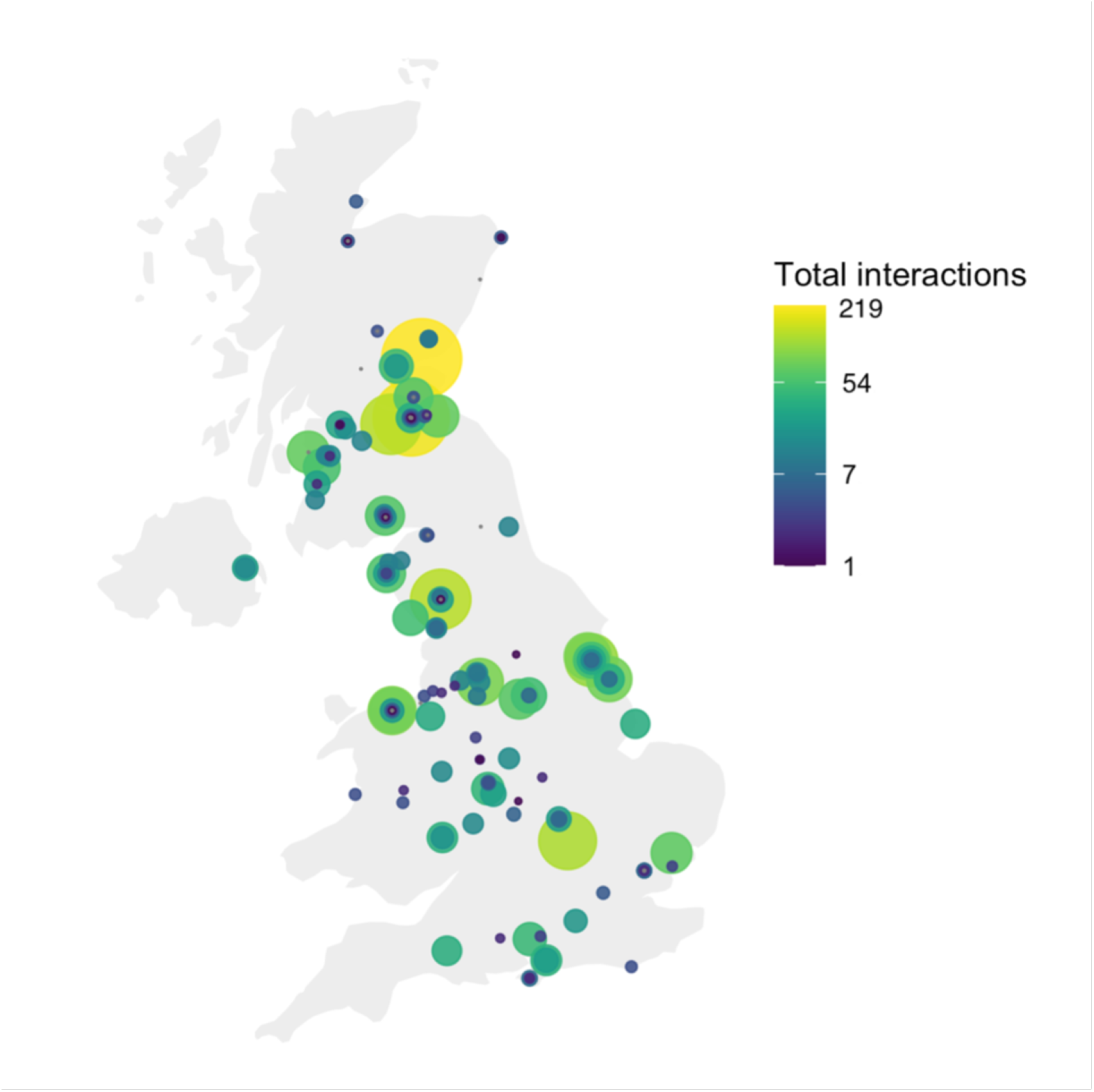
The number of responses from the areas that posted service change updates in the U.K.

### Strengths and limitations

This study demonstrated its strengths by extracting data from user-frequented sites that were appropriate for the research. However, due to the data protection laws of the NHS, we were unable to access information on emails, SMS, or letters addressed that were sent to patients regarding changes in the service.

### Future directions

This research is the first to evaluate social media sites to investigate notifications of changes to the audiology service. In future research, we will further extend this study by incorporating additional platforms, including other electronic media sites such as, TikTok and YouTube. To enhance our understanding of the communication strategies employed by healthcare providers, we may also consider supplementing social media-mined data with surveys sent to various National Health Service (NHS) trusts. Such surveys will help us to determine the approaches utilised by these trusts in informing patients of service changes and suggest potential areas for improvement to deliver a superior service.

## Conclusion

The current study investigated how several NHS audiology departments used social media to notify patients of service changes. It found that social media was not being used to its maximum potential by many trusts in the UK, as supported by the lack of posts or information on service changes on their Facebook sites and even less on Twitter. Audiology healthcare practitioners who work with patients with hearing loss, should be aware of how crucial information accessible via electronic media facilitates service accessibility for individuals with hearing loss. A well-informed workforce would provide more effective service and would in turn lead to fewer patient complaints. Social media has been utilised in the past for information dissemination and updates during public health crises [15].

## Data Availability

All relevant data are within the manuscript and its Supporting Information files.

## Acknowledgments

This research is supported by the UK EPSRC COG-MHEAR programme (Grant No. EP/M026981/1).

## References

1. Swanepoel DW, Hall JW. Making audiology work during COVID-19 and beyond. The Hearing Journal. 2020;73(6):20–22.

2. Parmar B, Beukes E, Rajasingam S. The impact of COVID-19 on provision of UK audiology services & on attitudes towards delivery of telehealth services. International Journal of Audiology. 2022;61(3):228–238.

3. Eikelboom RH, Swanepoel DW. International survey of audiologists’ attitudes toward telehealth. American Journal of Audiology. 2016;25(3S):295–298.

4. Saunders GH, Roughley A. Audiology in the time of COVID-19: practices and opinions of audiologists in the UK. International Journal of Audiology. 2021;60(4):255–262.

5. Manchaiah V, Bellon-Harn ML, Kelly-Campbell RJ, Beukes EW, Bailey A, Pyykkő I. Media use by older adults with hearing loss: An exploratory survey. American Journal of Audiology. 2020;29(2):218–225.

6. UK’s internet use surges to record levels; 2020. https://www.ofcom.org.uk/news-centre/2020/uk-internet-use-surges#:~:text=The%20proportion%20of%20UK%20adults,(from%2013%25).).

7. Gupta P, Khan A, Kumar A. Social media use by patients in health care: a scoping review. International Journal of Healthcare Management. 2022;15(2):121–131.

8. Lee JW, Bance ML. Hearing loss. Practical neurology. 2019;19(1):28–35.

9. Zimmer M. “But the data is already public”: on the ethics of research in Facebook. Ethics and information technology. 2010;12:313–325.

10. Hussain Z, Sheikh Z, Tahir A, Dashtipour K, Gogate M, Sheikh A, et al. Artificial Intelligence–Enabled Social Media Analysis for Pharmacovigilance of COVID-19 Vaccinations in the United Kingdom: Observational Study. JMIR Public Health and Surveillance. 2022;8(5):e32543.

11. Timeline of UK coronavirus lockdowns, March 2020 to March 2021; 2021. https://www.instituteforgovernment.org.uk/sites/default/files/timeline-lockdown-web.pdf.

12. Waterloo SF, Baumgartner SE, Peter J, Valkenburg PM. Norms of online expressions of emotion: Comparing Facebook, Twitter, Instagram, and WhatsApp. New media & society. 2018;20(5):1813–1831.

13. Barak A, Sadovsky Y. Internet use and personal empowerment of hearing-impaired adolescents. Computers in human behavior. 2008;24(5):1802–1815.

14. Pilling D, Barrett P. Text communication preferences of deaf people in the United Kingdom. Journal of deaf studies and deaf education. 2008;13(1):92–103.

15. Saroj A, Pal S. Use of social media in crisis management: A survey. International Journal of Disaster Risk Reduction. 2020;48:101584.

